# How appropriate is use of antiplatelet medications in patients with transient ischemic attacks and stroke: an analysis of 9132 patients over eight years

**DOI:** 10.1101/2023.02.20.23286209

**Authors:** Hiba Naveed, Naveed Akhtar, Salman Al-Jerdi, Ryan Ty Uy, Sujatha Joseph, Deborah Morgan, Blessy Babu, Shobana Shanthi, Ashfaq Shuaib

## Abstract

**Background and purpose:** Guidelines recommend patients with high-risk TIAs and minor stroke presenting within 1-3 days from onset be offered dual antiplatelet therapy (DAPT). There is little data on real-world adherence to these recommendations. We evaluated the appropriateness of DAPT use in TIA and stroke patients in a prospective Database.

**Methods:** The Qatar Stroke Database began enrollment of patients with TIAs and acute stroke in 2014 and currently has ∼ 16,000 patients. For this study we evaluated the rates of guideline-adherent use of antiplatelet treatment at the time of discharge in patients with TIAs and stroke. TIAs were considered high-risk with ABCD2 score of ⍰ 4 and minor stroke was defined as NIHSS ⍰ 3. Patient demographics, clinical features, risk factors, previous medications, imaging and laboratory investigations, final diagnosis, discharge medications, and discharge and 90-day modified Rankin Scale (mRS) were analyzed.

**Results:** After excluding patients with ICH, mimics and rare secondary causes, 8082 patients available for final analysis (TIAs: 1357;stroke 6725). In high-risk TIAs, 282 of 666 (42.3%) patients were discharged on DAPT. In patients with minor stroke, 1207 of 3572 (33.8%) patients were discharged on DAPT. DAPT was inappropriately offered to 238 of 691 (34.4%) of low-risk TIAs and 809 of 3153 (25.7%) of non-minor stroke patients.

**Conclusions:** This large database of prospectively collected patients with TIAs and stroke shows that, unfortunately, despite several guidelines, a large majority of patients with TIAs and stroke are receiving inappropriate antiplatelet treatment at discharge from hospital. This requires urgent attention and further investigation.

## Introduction

Early initiation of antithrombotic medications decreases the risk of recurrence of vascular events in patients presenting with transient ischemic attack (TIA) and ischemic stroke (1-2). Numerous clinical trials during the last 50 years have provided convincing evidence that medications such as ASA and clopidogrel reduce the risk of stroke and other vascular events when compared to placebo (3-4). Following the successful use of dual antiplatelet treatment (DAPT) in patients with acute coronary syndrome (5), there was excitement that similar long-term combination treatment may be superior to single antiplatelet therapy (SAPT) in patients with TIAs and ischemic stroke. This was initially not supported by clinical evidence following completion of the MATCH trial, where long-term use of ASA and clopidogrel was shown not to be superior to clopidogrel alone (6). The combination treatment was also associated with a higher risk of intracranial and systemic hemorrhage. DAPT was therefore abandoned as a long-term strategy in stroke prevention in most guidelines (7-8).

In the early 2000’s, studies from San Francisco (9), Oxfordshire (10) and Paris (11) reported that the risk of recurrent TIAs and stroke was high following a TIA if the patients are examined early following the ictal event. Also, importantly, the high recurrence risk was attenuated with rapid assessment and treatment (10-11). The studies also helped determine identification of ‘high-risk’ TIA patients in whom early recurrence was more pronounced (12). Thus, the age of the patient, presence of vascular risk factors (hypertension and diabetes), duration of focal deficits and the presence of motor symptoms were identified as important factors that increased the risk of early recurrence (9-12). The ABCD_2_ score incorporates these elements into an equation and is the most widely used score to determine stroke risk in patients with TIAs (13). More recently, imaging criteria has been added to the ABCD_2_ stroke-risk score (14) to help further improve the identification of patients in whom early recurrence is particularly high.

There was renewed interest in the short-term use of DAPT following the observations that the risk of recurrent stroke is very high in patients with ABCD_2_ score of ≥ 4. The small FASTER trial was the first to report in 2012 that short-term 90 days DAPT was superior to SAPT in patients with high-risk TIAs when treated within 24 hours of onset of symptoms (15). There were 392 patients recruited in the small study that showed a significantly lower stroke risk of 7.1% in the DAPT arm compared to 10.8% in the SAPT arm (15). Two larger more recent trials have shown that DAPT for 21-90 days initiated early following a high-risk TIA or minor stroke may result in a significant reduction of stroke recurrence (16-17). In the CHANCE trial, DAPT was used for 21 days in patients presenting within 24 hours from onset of symptoms (16), whereas the POINT trial used DAPT for 90 days in patients with high-risk TIAs or minor stroke (NIHSS ≤ 3 on admission), initiated within 12 hours from onset of symptoms (17). The significantly better outcome with combination ASA and clopidogrel treatment resulted in strong “level A class I” recommendation that DAPT be used in patients with high-risk TIAs and minor stroke if they present within 1-3 days from onset of symptoms, with the recommendation appearing in guideline statements from the USA (7-8), Canada (18), and Europe (19).

The recommendations for the use of DAPT are specific for high-risk TIAs (ABCD_2_ score of ≥ 4) or minor stroke (NIHSS ≤ 3 on admission). However, there is very little information offered in the guidelines on the avoidance or inappropriate use of DAPT in low risk TIAs or patients with moderate to large stroke (NIHSS of ≥ 4). Two recent reports published in 2022 show that DAPT was under-utilized in high-risk patients with minor stroke (20-21). The Get-With-The-Guidelines Stroke program, which evaluates stroke treatment in 1800 hospitals, reported that only 47% of patients with minor stroke were offered DAPT in 2019. The same study also showed that inappropriate DAPT was offered to 42.6% of patients with non-minor stroke (20). The second study from the same cohort showed that there was an increase in the use of DAPT following the publication of the CHANCE and POINT studies but that it remains very low. DAPT in high-risk patients was higher in comprehensive stroke centers (54.7%) compared to primary stroke centers (23.1%) (21).

The low rates of guideline adherence in primary and comprehensive centers in the USA is worrisome and requires confirmation from other prospectively acquired registries. In this communication, we report on the antiplatelet treatment in 8082 patients with TIAs (n=1357) and ischemic stroke (n=6725) who were entered prospectively in our stroke registry between 2013 and 2022). Our specific aims include evaluation of factors that were associated with low rates of DAPT usage in patients with high-risk TIAs and minor stroke. An additional goal was to identify the reasons for inappropriate use of DAPT in low-risk TIAs or patients with nonminor stroke.

## Methods

This study comprised of patients that were prospectively entered into the stroke program database. The study was approved by the Committee for Human Ethics Research, Academic Health Service at HMC (MRC-01-20-1135).

All patients admitted via the emergency department with a provisional diagnosis of stroke, including ischemic stroke, stroke mimics, transient ischemic attack (TIA) and intracranial hemorrhage (ICH) admitted to Hamad General Hospital (HGH), Doha, Qatar between January 1, 2014 through December 04, 2021 were entered into the database by four especially trained advanced nurse practitioners. For the purpose of this study, we only evaluated patients with a confirmed diagnosis of TIA and ischemic stroke. We excluded patients in whom the final diagnosis was either ICH or stroke mimics. The details of the stroke database have previously been published (22-27). In brief, HGH is a Joint Commission International accredited 600-bed hospital, where ∼95% of all strokes in Qatar requiring hospitalization are admitted. The stroke program, also certified by the Joint Commission International, is equipped with all the necessary laboratory, neuro-radiological and neurosurgical facilities needed to manage acute stroke patients and has 24-hour thrombolysis and thrombectomy services. The stroke service is run by 7 stroke-trained neurologists and includes clinical pharmacists, nurse-practitioners, physical and occupational therapists and speech-language pathologists. All patients with a suspected diagnosis of acute stroke are evaluated in the Emergency Department (ED) by the Stroke team to enable immediate decisions on acute stroke interventions and further management. The majority of patients are admitted to a designated stroke ward.

### A. Patient Characteristics and Data Collection

The anonymized data entered into the HGH Stroke Database (Microsoft Office Access 2007 Database) includes patient characteristics such as age, sex, nationality, medical comorbidities and prior medication. We specifically evaluated the prior use of antithrombotic medications and whether the patients were taking any medications for common stroke risk factors, including antihypertensive, antidiabetic and statin medications.

Upon identification in the ED, data was collected once confirmation of diagnosis of ischemic stroke was made using the *International Classification of Disease, 10th Edition*, definitions (H34·1, I63.x, I64.x, I61.x, I60.x, G45.x). Data from emergency medical services/paramedics, immediate ED care, door-to-needle time (for thrombolysis patients), prior mRS, NIHSS score, length of stay (LOS), neuroimaging, post-stroke complications, in-hospital mortality, and recurrences were recorded. The modified Rankin scale (mRS) measurements were done at discharge and at 90 days (28). Patients were classified as having a good (mRS of ≤0-2) or poor (mRS 3-6) outcome. We used the TOAST classification (29) for the final diagnosis of stroke etiology. This classification defines patients in the following categories with predefined criteria; large vessel disease, small vessel or lacunar stroke (SVD), cardioembolic, and stroke of determined or undetermined etiology (29).

Hypertension was diagnosed with a blood pressure higher than 140 mmHg systolic and 90 mmHg diastolic as defined by the International Society of Hypertension (31). Dyslipidemia was defined as a low-density lipoprotein-cholesterol (LDL) level ≥ 3.62 mmol/L, high-density lipoprotein-cholesterol (HDL) level ≤ 1.03 mmol/L, triglycerides ≥ 1.69 mmol/L, or current treatment with a cholesterol-lowering drug (32). Atrial Fibrillation (AF) was diagnosed based on electrocardiographic findings on admission or on Holter monitoring during hospitalization. Smoking was defined as current cigarette smoking. Complications monitored and recorded included aspiration pneumonia, urinary tract infection, bedsores and sepsis during hospitalization. Diabetes was diagnosed according to the American Diabetes Association (ADA) and WHO recommendations and included patients with a previous diagnosis of DM, on medication for DM or a HbA1c ≥6.5% and the diagnosis of pre-DM was based on a HbA1c of 5.7 - 6.4 % as per 2015 ADA clinical practice recommendations (33).

For this study, we specifically evaluated the timing of initiation of antiplatelet treatment in patients with non-cardioembolic TIAs and ischemic strokes. The prior use of any antiplatelet use was documented. We also documented the time from initial diagnosis of TIA or ischemic stroke and initiation of SAPT or DAPT treatment in hospital for all patients. The Database records the patient outcomes at 90 days (mRS) and one-year follow-up of major cardiovascular events (MACE) but unfortunately does not document if the patient develops any complications (specifically intracranial or systemic bleeding). We are therefore unable to determine bleeding related complications in the patients in the Database.

To collect post-discharge, follow up data, the Cerner electronic medical systems were used to track patient admissions throughout the state of Qatar. We coded patient outcome using major MACE scores based on data in the Cerner files. We collected data on recurrent stroke, post-stroke myocardial infarction (non-fatal and fatal), cardiac arrest, post-stroke cardiac revascularization and death for one year.

### B. Appropriate use of antiplatelet agents

The main objective of the current study was the appropriate use of antiplatelet agents in patients presenting with a diagnosis of TIA or acute stroke. We used the American Heart Association/American Stroke Association recommendation for DAPT use (21-90 days) in patients with high-risk TIA and minor stroke. Patients with TIAs within 3 days and ABCD_2_ score of ≥ 4 and NIHSS ≤ 3 on admission were considered appropriate for DAPT (8). We therefore analyzed the data in four major categories as shown in table 1. Patients with TIAs (n=1357) were classified into high-risk (n=666) and low-risk (691) TIAs. The appropriate use of DAPT and SAPT was evaluated in this group. We considered DAPT appropriate only in patients with ABCD_2_ score of ≥ 4 in whom treatment could be initiated within three days from symptoms onset. DAPT in all other TIA patients was considered inappropriate. For patients with completed stroke we considered the use of DAPT ‘appropriate’ when treatment was initiated within three days in patients with NIHSS ≤ 3. In all other patients with stroke the use of DAPT was considered ‘inappropriate’.

**Table 1.**
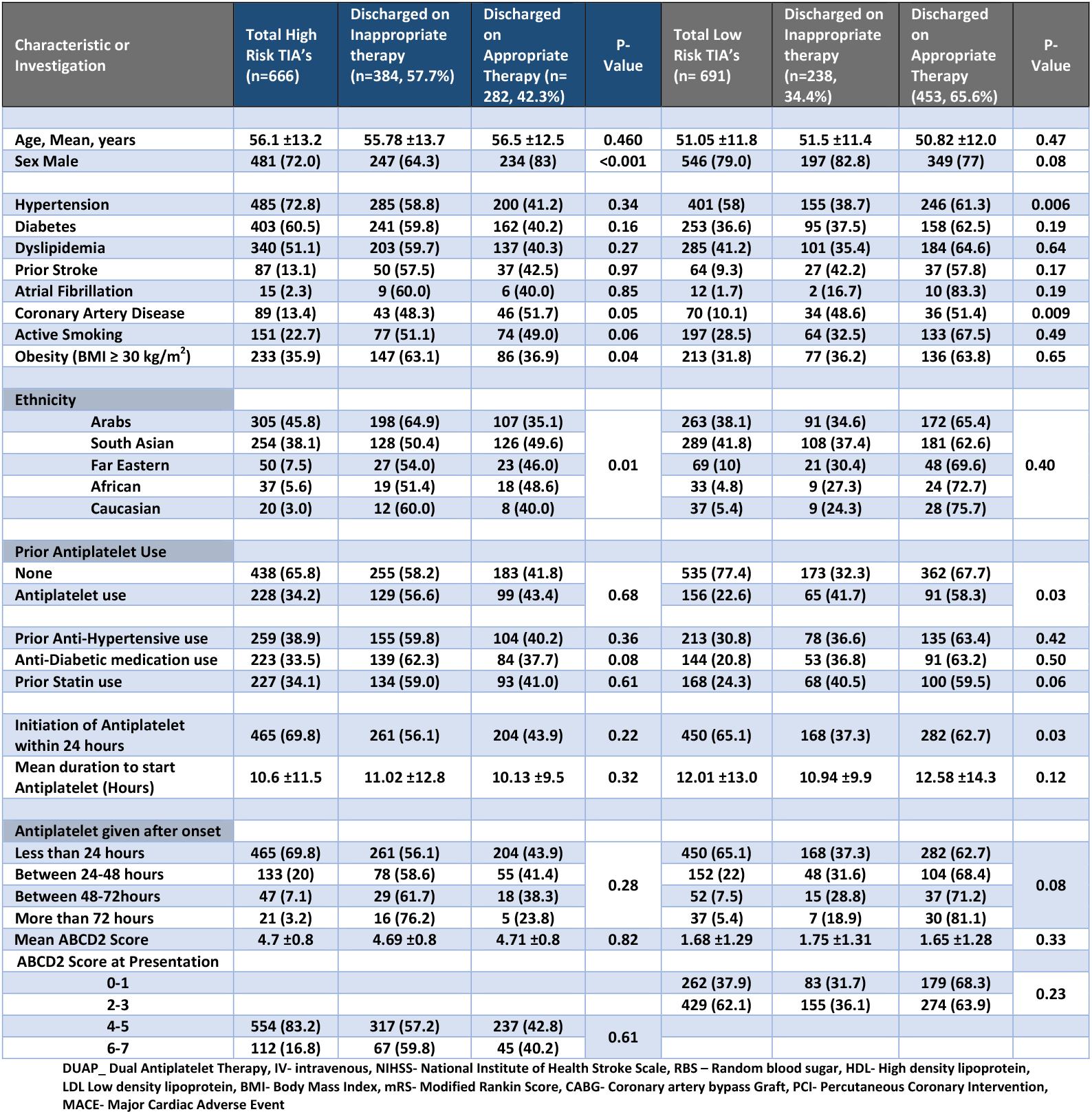
Baseline Characteristics of patients admitted with TIA and discharged on appropriate antiplatelet therapy.

Antiplatelet treatment was categorized into the following groups: ASA alone, clopidogrel alone, ASA=clopidogrel, ASA=dipyridamole, ASA=other antiplatelet agents such as ticlopidine, prasugrel, cilostazol or ticagrelor. Although DAPT may include any two antiplatelet agents, the combination of ASA=clopidogrel were the only one used in the Database.

### C. Data Analysis and Statistics

Descriptive results for all quantitative variables (e.g., age, systolic BP, BMI and others as shown in Table 1) were reported as mean ± standard deviation (SD). Numbers (percentage) were reported for all categorical variables (e.g., gender, diabetes, atrial fibrillation and others as shown in Table 1). The distribution of continuous variables was assessed by applying Kolmogorov Smirnov tests prior to using statistical tools.

One Way ANOVA was applied to test for significant differences across antiplatelet use for all continuous variables whereas; chi-square tests were used for all categorical variables. Independent sample t-test was used to compare the average for all the quantitative variables between patients with and without diabetes and Pearson Chi-Square test or Fisher Exact test was used to compare the proportion of all qualitative variables between patients with and without diabetes.

A multiple logistic regression analysis for the variables significant at univariate analysis was performed to determine if demographic (e.g., age, gender etc.) and clinical characteristics were associated with those who had mild stroke (NIHSS ≤=4) in comparison to moderate -severe strokes. The Wald test was computed on each predictor to determine which were significant. Adjusted Odds ratio and 95% confidence interval for the odds ratio were reported.

A p-value 0.05 (two tailed) was considered statistically significant level. SPSS 21·0 statistical package was utilized for the analysis.

## Results

During the study period (2014-2022), there were 15,993 patients prospectively enrolled in the Database. We excluded patients with ICH (1624), stroke mimics (4563), cerebral venous sinus thrombosis (214) and patients that required long-term anticoagulation (640), leaving us with a total of 8082 patients in whom discharge medications included single or dual antiplatelet medications. There were 1357 patients with TIAs (including 666 patients with ABCD2 score of ≥ 4) and 6725 patients with ischemic stroke (including 3572 patients with minor stroke [NIHSS ≤=4]. The patient population was predominantly male expatriate workers reflecting the demographics of Qatar that has a large expatriate working population (http://www.mdps.gov.qa/english/population). Female patients were mostly Qatari locals or long-stay expatriates and were significantly older in age. The expatriate patient population was mostly Arabs (45.8%) or South Asians (38.1%).

### A. Prior use of antiplatelet agents at the time of the TIA or stroke

Prior to the TIA or stroke 6291 (77.8%) of patients were *naive* to antiplatelet treatment. Aspirin was the most common antiplatelet agent (1304, 16.1%) prior to the acute event. Clopidogrel was the SAPT in 149, 1.8%) patients. Of the 1357 TIA patients, 322 (23.7%) were on SAPT and only 62 (4.6%) were taking DAPT. There was no difference in the appropriate or inappropriate use of DAPT in relationship to the prior use of antiplatelet therapy as shown in table 1. Similarly, the majority of patients with stroke were also not on any antiplatelet treatment prior to the acute event (5318 of 6725 [79.1%]. There were 1131 (16.8%) on prior SAPT at the time of the stroke, including 17% with minor stroke and 16.5% with nonminor stroke.

### B. Distribution of antiplatelet treatment in the patients with TIAs

The distribution of antiplatelet treatment at the time of discharge for TIAs and minor stroke is shown in table 1A. In patients with high-risk TIAs, 282/666 [42.3%] were discharge on DAPT and 384/666 [57.7%] on SAPT. At the time of discharge, SAPT was offered to 453/691 [65.6%] of low-ABCD_2_ patients and DAPT offered to 238/691 [34.4%] such patients. We next evaluated the relationship between increasing ABCD2 scores and usage of DAPT. As shown in table 1, there appeared to be no difference in the DAPT use with ABCD2 score between 4 and 7. The frequency of DAPT use in patients with ABCD_2_ score of 0-3 was marginally lower, especially in the ABCD2 with scores of 0-2. In our population, there is a higher frequency of symptoms related to the posterior circulation. There were 274/1357 (20.2%) of TIA’s of posterior circulation origin and 55% had ABCD2 score of ≤3. The recommended treatment in such patients is SAPT as the CHANCE (16) and POINT (17) trials excluded these patients and the most current national guidelines (7, 8, 18-19) do not recommend DAPT in such patients.

We next evaluated if patients presenting within 24 hours from onset were more likely to be offered DAPT. As shown in table 1, time to treatment from onset seems to make no difference in treatment. The rates of DAPT were similar in patients presenting early (within 24 hours) or late (more than 72 hours).

We also evaluate whether the prior use of antiplatelet agents may influence DAPT following a TIA. As shown in table 1, most patients with TIAs were naïve to antiplatelet agents at the time of the acute event. There was no difference in the frequency of DAPT in patients with or without previous ASA use in patients with low or high-risk categories. Very few patients were on DAPT prior to the presentation. In these patients the rates of DAPT at the discharge was significantly higher when compared the other two groups. Other factors, including the relationship of DAPT to preexisting vascular risk factors and ethnicity of the patients is shown in table 1.

### C. Distribution of antiplatelet treatment in patients with ischemic stroke

This group comprised 6725 patients. As shown in table 2, 53.1% of patients with acute stroke had an NIHSS score of ≤=3. In patients with minor stroke, 1207/3572 [33.8%] patients were discharged on DAPT and 2365/3572 (66.2%) on SAPT. For patients with NIHSS of ≥4, SAPT is recommended for antiplatelet therapy by most international guidelines (7,8,18-19). In our patients, SAPT was offered to 2344/3153 [74.3%] patients and DAPT was offered to 809/3153 [25.7%] patients. As shown in figure 2, the rates of DAPT use decreased as the NIHSS increased. The rates of DAPT use was similar in patients presenting within <24 hours or more than 72 hours from onset of symptoms.

**Table 2.** Baseline Characteristics of patients admitted with Ischemic Stroke and discharged on appropriate therapy.

| Characteristic or Investigation | All Minor Strokes (n= 3572) | Discharged on Inappropriate therapy (n= 2365, 66.2%) | Discharged on Appropriate Therapy (n= 1207, 33.8%) | P- Value | All Major Strokes (n= 3153) | Not Discharged on Appropriate therapy (n= 809, 25.7%) | Discharged on Appropriate Therapy (2344, 74.3%) | P-Value |
| --- | --- | --- | --- | --- | --- | --- | --- | --- |
| Age, Mean, years | 54 ±12.3 | 54.6 ±12.7 | 54.3 ±11.5 | 0.43 | 55.18 ±37.4 | 54.1 ±11.7 | 55.5 ±42.9 | 0.36 |
| Male Sex | 2899 (81.2) | 1855 (78.4) | 1044 (86.5) | <0.001 | 2606 (82.7) | 702 (86.8) | 1904 (81.2) | <0.001 |
| Hypertension | 2672 (74.8) | 1744 (65.3) | 928 (34.7) | 0.04 | 2264 (71.8) | 620 (27.4) | 1644 (72.6) | <0.001 |
| Diabetes | 2040 (57.1) | 1311 (64.3) | 729 (35.7) | 0.005 | 1784 (56.6) | 487 (27.3) | 1297 (72.7) | 0.02 |
| Dyslipidemia | 1778 (49.8) | 1144 (64.3) | 634 (35.7) | 0.02 | 1441 (45.7) | 395 (27.4) | 1046 (72.6) | 0.04 |
| Prior Stroke | 374 (10.5) | 226 (60.4) | 148 (39.6) | 0.01 | 380(12.1) | 121 (31.8) | 259 (68.2) | 0.004 |
| Atrial Fibrillation | 81 (2.3) | 63 (77.8) | 18 (22.2) | 0.03 | 133 (4.2) | 20 (15.0) | 113 (85.0) | 0.004 |
| Coronary Artery Disease | 354 (9.9) | 190 (53.7) | 164 (46.3) | <0.001 | 356 (11.3) | 123 (34.6) | 233 (65.4) | <0.001 |
| Active Smoking | 1048 (29.3) | 673 (64.2) | 375 (35.8) | 0.11 | 945 (30) | 265 (28.0) | 680 (72.0) | 0.04 |
| Obesity (BMI ≥ 30 kg/m<sup>2</sup>) | 919 (26.3) | 627 (68.2) | 292 (31.8) | 0.09 | 753 (24.8) | 172 (22.8) | 581 (77.2) | 0.02 |
| Ethnicity |  |  |  |  |  |  |  |  |
| Arabs | 1197 (33.5) | 812 (67.8) | 385 (32.2) | 0.29 | 924 (29.3) | 218 (23.6) | 706 (76.4) | 0.38 |
| South Asian | 1842 (51.6) | 1199 (65.1) | 643 (34.9) |  | 1767 (56) | 468 (26.5) | 1299 (73.5) |  |
| Far Eastern | 309 (8.7) | 197 (63.8) | 112 (36.2) |  | 259 (8.2) | 73 (28.2) | 186 (71.8) |  |
| African | 140 (3.9) | 97 (69.3) | 43 (30.7) |  | 134 (4.2) | 31 (23.1) | 103 (76.9) |  |
| Caucasian | 84 (2.4) | 60 (71.4) | 24 (28.6) |  | 69 (2.2) | 19 (27.5) | 50 (72.5) |  |
| Prior Antiplatelet Use |  |  |  |  |  |  |  |  |
| None | 2808 (78.6) | 1887 (67.2) | 921 (32.8) | <0.02 | 2510 (79.6) | 615 (24.5) | 1895 (75.5) | <0.003 |
| Antiplatelet use | 764 (21.4) | 478 (62.6) | 286 (37.4) |  | 643 (20.4) | 194 (30.2) | 449 (69.8) |  |
| Prior Anti-Hypertensive use | 1238 (34.7) | 851 (68.7) | 387 (31.3) | 0.02 | 918 (29.1) | 245 (26.7) | 673 (73.3) | 0.39 |
| Anti-Diabetic medication use | 1055 (29.5) | 700 (66.4) | 355 (33.6) | 0.91 | 738 (23.4) | 213 (28.9) | 525 (71.1) | 0.02 |
| Prior Statin use | 829 (23.2) | 540 (65.1) | 289 (34.9) | 0.45 | 659 (20.9) | 184 (27.9) | 475 (72.1) | 0.13 |
| Initiation of Antiplatelet within 24 hours | 1610 (45.1) | 985 (61.2) | 625 (38.8) | <0.001 | 1386 (44.0) | 387 (27.9) | 999 (72.1) | <0.01 |
| Mean duration to start Antiplatelet (Hours) | 13.5 ±17.7 | 14.2 ±20.3 | 12.03 ±11.2 | 0.0001 | 16.8 ±20.9 | 14.9 ±22.6 | 17.5 ±20.2 | 0.003 |
| Antiplatelet given from Onset |  |  |  |  |  |  |  |  |
| Less than 24 hours | 1610 (45.1) | 985 (61.2) | 625 (38.8) | <0.001 | 1386 (44) | 387 (27.9) | 999 (72.1) | <0.001 |
| Between 24-48 hours | 882 (24.7) | 616 (69.8) | 266 (30.2) |  | 1179 (37.4) | 259 (22.0) | 920 (78.0) |  |
| Between 48-72 hours | 592 (16.6) | 391 (66.0) | 201 (34.0) |  | 362 (11.5) | 118 (32.6) | 244 (67.4) |  |
| More than 72 hours | 488 (13.7) | 373 (76.4) | 115 (23.6) |  | 226 (7.2) | 45 (19.9) | 181 (80.1) |  |
| NIHSS on admission (mean) | 1.5 ±1.1 | 1.54 ±1.1 | 1.61 ±1.1 | 0.08 | 8.61 ±5.5 | 7.50 ±4.6 | 9 ±5.7 | 0.0001 |
| NIHSS Admission |  |  |  |  |  |  |  |  |
| 0 | 853 (23.9) | 578 (67.8) | 275 (32.2) | 0.32 |  |  |  |  |
| 1 | 740 (20.7) | 496 (67.0) | 244 (33.0) |  |  |  |  |  |
| 2 | 1096 (30.7) | 728 (66.4) | 368 (33.6) |  |  |  |  |  |
| 3 | 883 (24.7) | 563 (63.8) | 320 (36.2) |  |  |  |  |  |
| 4 |  |  |  |  | 717 (31.0) | 227 (31.7) | 490 (68.3) | 0.04 |
| 5 |  |  |  |  | 511 (22.1) | 152 (29.7) | 359 (70.3) |  |
| 6 |  |  |  |  | 368 (15.9) | 105 (28.5) | 263 (71.5) |  |
| 7 |  |  |  |  | 239 (10.3) | 61 (25.5) | 178 (74.5) |  |
| 8 |  |  |  |  | 217 (9.4) | 58 (26.7) | 159 (73.3) |  |
| 9 |  |  |  |  | 132 (5.7) | 26 (19.7) | 106 (80.3) |  |
| 10 |  |  |  |  | 128 (5.5) | 28 (21.9) | 100 (78.1) |  |

| TOAST Classification |  |  |  |  |  |  |  |  |
| --- | --- | --- | --- | --- | --- | --- | --- | --- |
| Small Vessel Disease | 2126 (59.6) | 1405 (66.1) | 721 (33.9) | <0.001 | 1308 (41.6) | 327 (25.0) | 981 (75.0) | <0.001 |
| Large Vessel Disease | 575 (16.1) | 329 (57.2) | 246 (42.8) |  | 848 (26.9) | 279 (32.9) | 569 (67.1) |  |
| Cardioembolic | 478 (13.4) | 353 (73.8) | 125 (26.2) |  | 571 (18.1) | 121 (21.2) | 450 (78.8) |  |
| Stroke of Determined Origin | 207 (5.8) | 134 (64.7) | 73 (35.3) |  | 278 (8.8) | 60 (21.6) | 218 (78.4) |  |
| Stroke of Undetermined Origin | 179 (5.0) | 138 (77.1) | 41 (22.9) |  | 143 (4.5) | 21 (14.7) | 122 (85.3) |  |

DAPT\_ Dual Antiplatelet Therapy, IV- intravenous, NIHSS- National Institute of Health Stroke Scale, RBS – Random blood sugar, HDL- High density lipoprotein, LDL Low density lipoprotein, BMI- Body Mass Index, mRS- Modified Rankin Score, CABG- Coronary artery bypass Graft, PCI- Percutaneous Coronary Intervention, MACE- Major Cardiac Adverse Event

**Figure 1:**
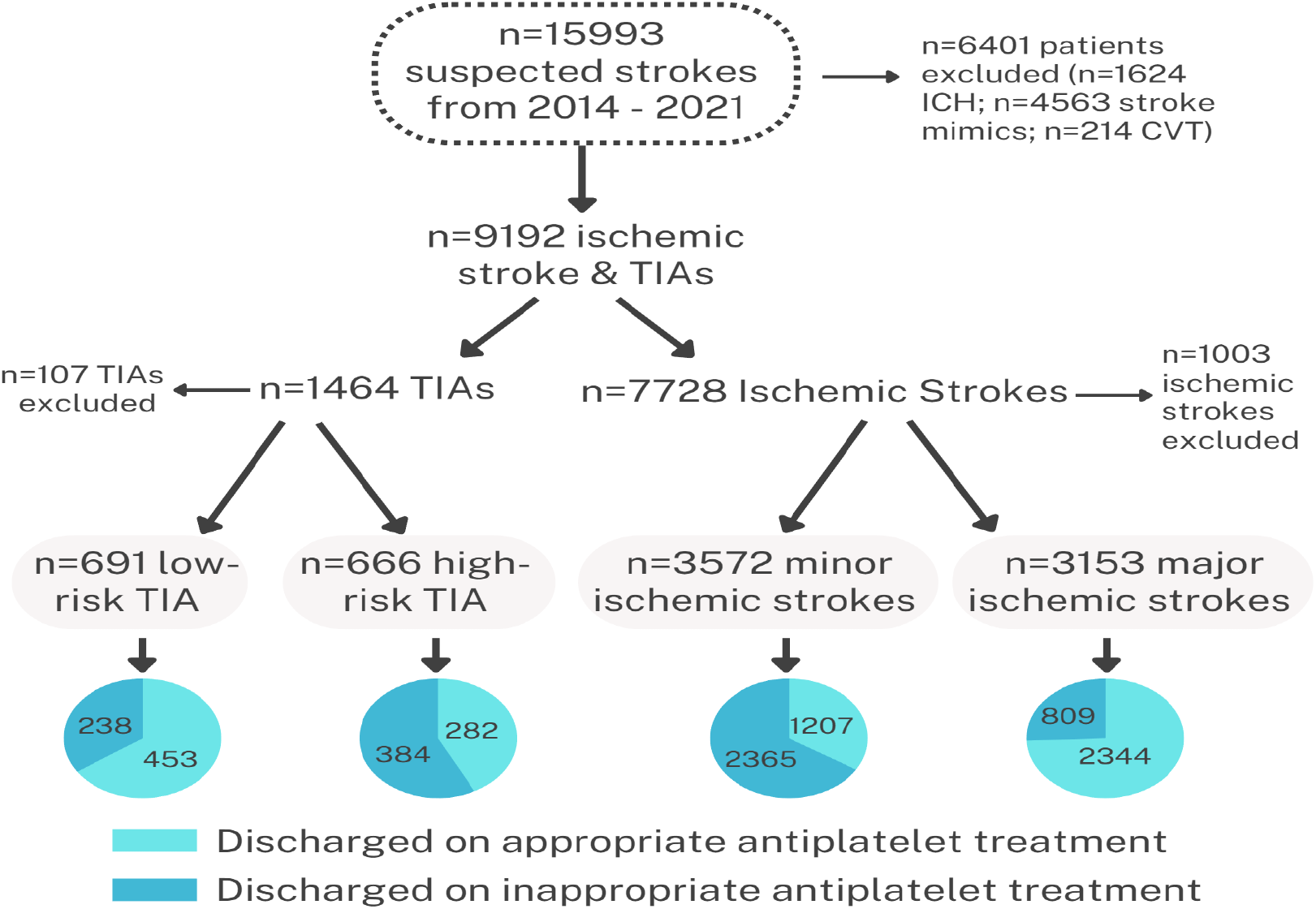
The flow diagram of the patients in the database used for the current analysis showing the total number of patients enrolled in the registry and the treatment at discharge in patients with TIAs and stroke

**Figure 2:**
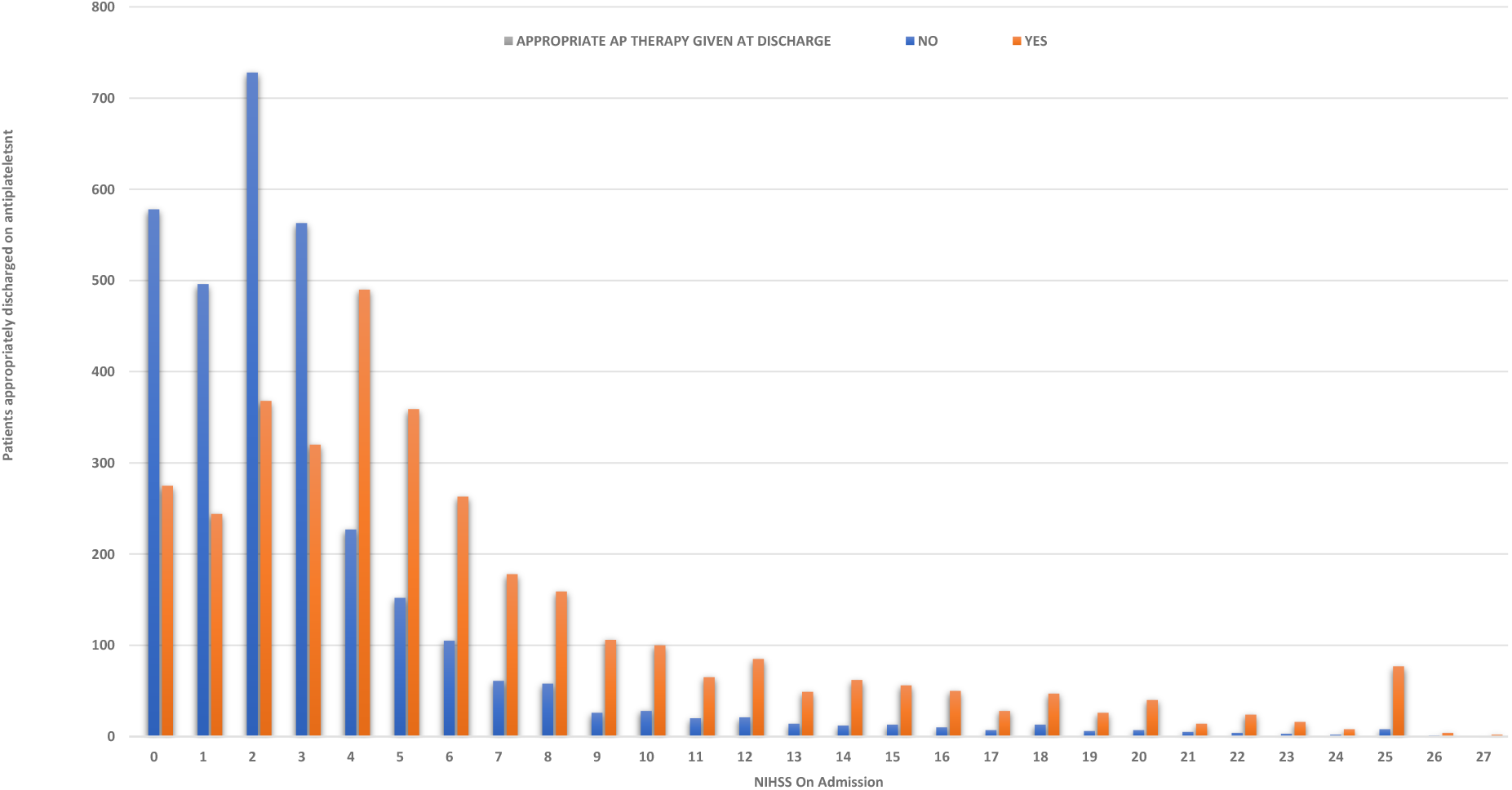
The treatment of DAPT in patients with acute stroke in relationship to the NIHSS

The relationship between vascular risk factors and the rates of DAPT used is shown in table 2. There were no differences in the use of DAPT in relationship to the prior use of antihypertensive, antidiabetic and statin medications in patients with acute stroke.

We used the TOAST classification for the underlying diagnosis of the acute stroke. As shown in table 2B, DAPT use was highest in minor strokes related to large vessel disease. DAPT was used in 246/575 [42.8%] in this high-risk group.

### D. Trends in DAPT use

We next explored some potential reasons for the low use of DAPT in our cohort. We initially evaluate whether there was a difference in the use of antiplatelet agents amongst the stroke consultants directly responsible for the care of the patients. In all there were 13 physicians that were responsible for the care of the stroke patients. There were no differences seen in the appropriate or inappropriate use of DAPT across the physician group (data not shown). Secondly, we evaluated the treatment trends across years of the study (2014 to 2022) and in the period before the CHANCE study (published in 2014), following the publication of CHANCE and POINT study (published in 2018) and the post-POINT time period. We were not able to show any significant increase in the DAPT use over time or following publication of the landmark papers (data not shown).

## Discussion

Antiplatelet medications are important preventive treatments in patients with TIAs and acute stroke (33-34). The risk reduction of recurrent vascular events with ASA alone is approximately 12 percent (35). The addition of clopidogrel may increase the risk reduction to 20 percent, (36) but this comes with an increased risk of hemorrhagic complications (36). Early initiation of DAPT within 24 hours of onset of symptoms in patients with high-risk TIAs and minor stroke has been shown to be superior to SAPT in preventing early recurrent stroke (16,17). Furthermore, recent sub-analysis of the CHANCE and POINT studies has shown that the risk of recurrent stroke is highest in the initial three days and later decreases rapidly (37-38). There is, therefore, consensus and guideline recommendations that DAPT use be restricted to high-TIAs and minor stroke patients when seen within 72 hours from onset of symptoms (7-8, 18-19). Research determining if the clinical practice has changed following the publications of the guidelines is important. Data from The-Get-With-The-Guidelines has shown that only 47 percent of patients with minor stroke were being offered DAPT. There was also an inappropriate 42.6% use of DAPT to patients with nonminor strokes (20-21). To our knowledge, there are no studies for appropriate use of DAPT in patients with stroke from other regions of the world. Similar studies on the appropriateness of antiplatelet medications in patients with TIAs have also not been published.

Our study shows that DAPT continues to be underutilized in patients presenting with high-risk TIAs, minor stroke and possibly inappropriately used in nonminor stroke. Only 42.3% of patients with TIAs and 33.8% of patients with minor stroke in our cohort were treated appropriately with DAPT at the time of discharge. DAPT was inappropriately used in 34.4% of low-risk TIAs and 25.7% of nonminor stroke patients. These numbers are similar to the report from the USA (20-21). We were unable to show any increase in the rates of DAPT use over time as the POINT trial was reported and the guidelines were revised. We also did not show any increase in the use of DAPT in patients who were on ASA at the time of the new event or in patients with higher vascular risk factors.

A number of issues require further discussion, including the methods used to identify high-risk patients, determination of the risks/benefits of inappropriate DAPT (especially in low-risk TIAs and in patients with nonminor stroke), recommendations for DAPT when the patients are already on ASA or are unable to take ASA because of a recent GI hemorrhage or allergy or have CYP2C19 loss of function.

Rapid identification of patients at high risk for recurrence is the most important initial step prior to initiation of appropriate antithrombotic treatment. The ABCD_2_ score is perhaps the most widely used score to identify high-risk TIA patients (13). A score of ≥ 4 is considered ‘high-risk’ and has been used to enter patients into clinical trials (16,17). In a recent meta-analysis of 29 studies, the 7-day risk of stroke was 10.2% in patients with an ABCD2 score of ≥ 4. A significantly lower risk of stroke of 3.2% was evident with lower ABCD2 scores (39). The meta-analysis also showed that while the higher scores were sensitive but not very specific for identifying patients at high early stroke risk (40), the addition of imaging to the ABCD2 score may improve its specificity (14).

There are additional clues from the recent large trials that identify TIA and minor strokes where DAPT is highly effective. In the CHANCE study, most strokes in both arms of the study occurred within the first two weeks of the symptom onset (45). Similarly, the POINT study also showed that recurrent stroke was most common in the initial three days and DAPT was likely to be effective if initiated within 3 days from onset of symptoms (46). Additionally, in both the POINT and CHANCE trials, patients with high blood pressure at the time of enrollment received a greater benefit from DAPT (47,48). The CHANCE trial also showed that increasing NIHSS, poor control of hypertension during follow-up, lack of use of lipid-lowering agents and the presence of intracranial atherosclerosis were additional risk factors for a higher risk of stroke recurrence (48).

The NIHSS score is used to define patients with minor stroke for the stroke prevention trials. An NIHSS score of **≤**=4 was used for identification of minor stroke in the two recent large DAPT studies (16,17). This arbitrary separation of minor from nonminor stroke is problematic as there are no biological reasons to expect a difference in outcome. It is therefore understandable that there is no clear separation of the use of DAPT and SAPT in patients when the NIHSS score increases beyond 4. Our results in a large cohort shows that DAPT use gradually decreases as the NIHSS rises but still remains high with NIHSS of 10. Almost 30% of patients were prescribed DAPT in the 5-10 NIHSS range. We don’t know if the risk of complications from DAPT rises with high NIHSS. In a small study of 119 patients with stroke and TIAs, bleeding risk increased with increasing NIHSS but this did not lead to a higher mortality (42). Another study presented at the American Academy of Neurology reviewed their results in 1316 patients in a prospectively collected database. The risk of cerebral hemorrhage was related to the size of the cerebral infarction and not to the single or dual antiplatelet treatment (43). Kim at al presented their data from a larger national registry in South Korea that included 4461 patients with moderate to large stroke (NIHSS 44-15). Fifty-two percent of patients were treated with SAPT and 47 percent received DAPT. During a three-month follow-up there were no significant differences in the recurrent stroke rates but the mortality was significantly lower in the DAPT group. The bleeding risk with the two therapies was however not reported (44). The limited available evidence suggests that DAPT is likely safe in patients with nonminor stroke.

We were not able to determine the reasons for the low use of DAPT in high-risk TIA patients. The prescribing patterns were similar for all the consultant neurologists and we did not see any increase in the use of DAPT with the availability of POINT study or with the wide availability of the revised guidelines. The emergency department is usually the first point of contact for the patient to be managed by the medical team. In a recent survey on the use of DAPT by emergency physicians in the USA, 68% of participants reported not using clinical prediction rules and only 18% reported the use of ABCD2 score to inform treatment choices (41). Nearly half the surveyed physicians reported that they will use ASA and more alarmingly, only 2% reported that they will treat high-risk TIA or minor stroke with DAPT (41). These are somber results and an opportunity for improving early emergency care of patients at the highest risk for recurrence.

There are several reasons for undertreatment with DAPT in high-risk patients. Perhaps the most difficult one to understand is the reluctance on the part of physicians to offer appropriate therapy. In our study, we found no differences in the use of DAPT between the consultants involved in the care of the patients. The Get-With-The-Guidelines program however showed remarkable differences between DAPT use in comprehensive centers (55.2%) versus primary stroke centers (22.3%) in the management of minor stroke (21). Other factors that contribute to the lower use of DAPT include the occurrence of a stroke while the patient is already on ASA (49), allergy to ASA (50), recent hemorrhage while on ASA (51) and perceived or known drug resistance to ASA or clopidogrel (51). It is not uncommon to have a vascular event while on ASA. In our series, approximately 20% of patients were on ASA at the time of their TIA or stroke. The recommended best approach is to add clopidogrel instead of switching to another agent (51). Allergy to ASA is rare and investigating for resistance is generally not recommend (51). Pseudo-resistance to ASA may be more common and may be related to enteric coating (51). It is important to emphasize that in the POINT trial, the overall risk of bleeding was low. The risk of major hemorrhage was 0.2% in the SAPT arm and 0.9% in the DAPT (53).

There are limitations to our study. Although the data was entered prospectively, this is a retrospective analysis and does not document the reasons DAPT or SAPT were chosen for individual patients. Although we recorded a large number of variables, we did not record specific complications to individual medications and therefore we are unable to document bleeding outcomes for the patients. Also we specifically did not look for symptomatic intracranial stenosis patients in whom arguably dual antiplatelets is recommended. This data is limited to antiplatelet medication use in a large comprehensive stroke program in the Middle East and the data may not be generalizable to other populations.

In summary, we reviewed the antiplatelet treatment patterns of TIAs and strokes during the last 8 years. During this time, especially following the publication of two important trials on the short-term use of DAPT in specific high-risk populations, a minority of patients were discharge on the appropriate prevention medications. More work at enhancing evidence-based adaptation of DAPT in the appropriate patients should be targeted for quality improvement initiatives.

## Data Availability

All data are available in the document. No additional data available

## Declaration of Competing Interests

None

### Ethical approval

The study was approved by the Institutional Review Board, Hamad Medical Corporation at the Medical Research Centre (MRC-01-20-1135) and the Institutional Review Board of Weill Cornell Medicine-Qatar (1932095-1/22-00016).

### Informed Consent

This is a registry study and hence consent Is not applicable

### Guarantor

Dr. Ashfaq Shuaib, also the corresponding author.

### Data availability statement

No additional data available

### Informed consent

Not applicable

### Contributorship

#### Concept, design, and draft

Hiba Naveed, Naveed Akhtar and Ashfaq Shuaib

#### Acquisition, analysis, interpretation of data, technical and administrative support

Hiba Naveed, Blessy, Deborah, Sujatha, Ryan, Shobana, Naveed Akhtar, Ashfaq Shuaib **Critical review:** Saadat Kamran, Salman Al-Jerdi, Ashfaq Shuaib

### Statistical analysis

Naveed Akhtar

## Acknowledgments

We acknowledge the assistance of all involved physicians, nurses, and staff of the Stroke Team. We also thank Ms. Reny Francis (HMC) for her editorial assistance and supportive care.

